# Spatiotemporal dissemination pattern of SARS-CoV-2 B1.1.28-derived lineages introduced into Uruguay across its southeastern border with Brazil

**DOI:** 10.1101/2021.07.05.21259760

**Authors:** Natalia Rego, Tamara Fernández-Calero, Ighor Arantes, Verónica Noya, Daiana Mir, Mariana Brandes, Juan Zanetti, Mailen Arleo, Emiliano Pereira, Tania Possi, Odhille Chappos, Lucia Bilbao, Natalia Reyes, Melissa Duquía, Matías Victoria, Pía Techera, María José Benítez-Galeano, Luciana Griffero, Mauricio Méndez, Belén González, Pablo Smircich, Rodney Colina, Cecilia Alonso, Gonzalo Bello, Lucía Spangenberg

## Abstract

During the first nine months of the SARS-CoV-2 pandemic, Uruguay successfully kept it under control, even when our previous studies support a recurrent viral flux across the Uruguayan-Brazilian border that sourced several local outbreaks in Uruguay. However, towards the end of 2020, a remarkable exponential growth was observed and the TETRIS strategy was lost. Here, we aimed to understand the factors that fueled SARS-CoV-2 viral dynamics during the first epidemic wave in the country. We recovered 84 whole viral genomes from patients diagnosed between November, 2020 and February, 2021 in Rocha, a sentinel eastern Uruguayan department bordering Brazil. The lineage B.1.1.28 was the most prevalent in Rocha during November-December 2020, P.2 became the dominant one during January-February 2021, while the first P.1 sequences corresponds to February, 2021. The lineage replacement process agrees with that observed in several Brazilian states, including Rio Grande do Sul (RS). We observed a one to three month delay between the appearance of P.2 and P.1 in RS and their subsequent detection in Rocha. The phylogenetic analysis detected two B.1.1.28 and one P.2 main Uruguayan SARS-CoV-2 clades, introduced from the southern and southeastern Brazilian regions into Rocha between early November and mid December, 2020. One synonymous mutation distinguishes the sequences of the main B.1.1.28 clade in Rocha from those widely distributed in RS. The minor B.1.1.28 cluster, distinguished by several mutations, harbours non-synonymous changes in the Spike protein: Q675H and Q677H, so far not concurrently reported. The convergent appearance of S:Q677H in different viral lineages and its proximity to the S1/S2 cleavage site raise concerns about its functional relevance. The observed S:E484K-VOI P.2 partial replacement of previously circulating lineages in Rocha might have increased transmissibility as suggested by the significant decrease in Ct values. Our study emphasizes the impact of Brazilian SARS-CoV-2 epidemics in Uruguay and the need of reinforcing real-time genomic surveillance on specific Uruguayan border locations, as one of the key elements for achieving long-term COVID-19 epidemic control.

## INTRODUCTION

By the end of 2020 and the beginning of 2021, several studies reported the emergence of novel SARS-CoV-2 variants of interest (VOI) and concern (VOC) with different missense mutations and deletions in the Spike (S) protein that impact viral transmissibility and escape from previous host’s immune responses, among other features. In Brazil, the SARS-CoV-2 lineages B.1.1.28 and B.1.1.33 dominated the first epidemic wave [1,2], but were replaced by the VOC P.1 and the VOI P.2, both descendants of lineage B.1.1.28 by the end of 2020 and beginning of 2021 (http://www.genomahcov.fiocruz.br/). The VOC P.1, which harbors the mutations of concern S:K417T/E484K/N501Y among its lineage defining mutations, was first detected in January 2021 [3]. Once originated in Manaus, capital of the Amazonas state in mid-November, rapidly spread across Brazil [4,5] and to over 50 countries globally (https://cov-lineages.org/lineage_description_list.html). The VOI P.2, carrying the concerning S:E484K amino acid change, was initially detected in samples from the Rio de Janeiro (RJ) state in October 2020 and spread throughout the country and abroad since then (https://cov-lineages.org/lineage_description_list.html) [6-8].

Uruguay was able to control the early viral dissemination during the first nine months of the SARS-CoV-2 pandemic. The low number of total cases, successful contact tracing, contained isolated outbreaks, and few deaths (https://guiad-covid.github.io/estadisticasuy.html) [9], were characteristic for this first period. At the beginning, viral diversity was high, with circulation of strains A.2, A.5, B.1, B.1.195 and B.31, introduced mostly through Montevideo, Uruguay’s capital city and connection hub by its international airport and harbor [10]. After multiple introductions of SARS-CoV-2 Brazilian lineages B.1.1.28 and B.1.1.33 into Uruguay mainly by the 1,068 km long Uruguayan–Brazilian dry border, these lineages were successfully disseminated becoming predominant since May, 2020 [11]. Towards the end of 2020, the number of active cases exponentially increased, from an average of 60 cases per day during October and November to more than 400 during December (https://guiad-covid.github.io/estadisticasuy.html), concomitant with the loss of the TETRIS (Test, Trace and Isolation strategy) safety zone [12,13]. SARS-CoV-2 positive daily new cases decreased after more stringent mobility measures were taken by the government during December 2020 (https://medios.presidencia.gub.uy/tav_portal/2020/noticias/AH_204/Medidas%2016.12.2020.pdf) [9], but the total number of cases stayed high outside the TETRIS zone. While a subsequent exponential growth period is characterized by the introduction and dissemination of the VOC P.1 from Brazil [14], there is a gap in knowledge concerning the factors that fueled SARS-CoV-2 viral dynamics during the first exponential increase of COVID-19 through the end of 2020 and beginning of 2021.

The VOI P.2 reached the southernmost Brazilian state of Rio Grande do Sul (RS) in October, 2020 and then spread massively in that region [15-18]. Given the high viral flux between RS and Uruguay detected during 2020 [11], it is likely that the introduction of the VOI P.2 from Brazil could have been associated with the epidemic worsening in Uruguay by the end of 2020 and beginning of 2021. To test this hypothesis, we conducted an epidemiological and genomic analysis of SARS-CoV-2 complete genomes from 84 patients. All patients were diagnosed between November 2020 and February 2021 in Rocha, a sentinel Eastern Uruguayan department bordering with RS, Brazil, which nearly replicates the epidemiological curve of the whole country (see P7/P14 calculations in https://guiad-covid.github.io/estadisticasuy.html). Our study revealed that lineage B.1.1.28 was the most prevalent viral SARS-CoV-2 variant in Rocha up to December, 2020, but it was mostly replaced by lineage P.2 in January, 2021.

## MATERIALS AND METHODS

### SARS-CoV-2 samples and ethical aspects

A total of 84 SARS-CoV-2 whole-genomes were recovered from nasopharyngeal-throat combined swabs samples collected from clinically ill or asymptomatic individuals that reside in Rocha, a southeast Uruguayan department at the Brazilian border (Figure 1A and Table S1). Additionally, two samples were collected in Artigas and Rivera, two departments in the Uruguayan north region that also border Brazil (Table S1). All samples were collected between November 25, 2020, and February 26, 2021, and underwent testing at Sanatorio Americano Montevideo (SASA, Molecular Biology Lab) and Universidad de la República (UdelaR) CURE Este, Rocha (Molecular Ecology Lab). Sequencing of the Uruguayan samples was held at Institut Pasteur de Montevideo (IPMON, Bioinformatics Unit).

**Figure 1.**
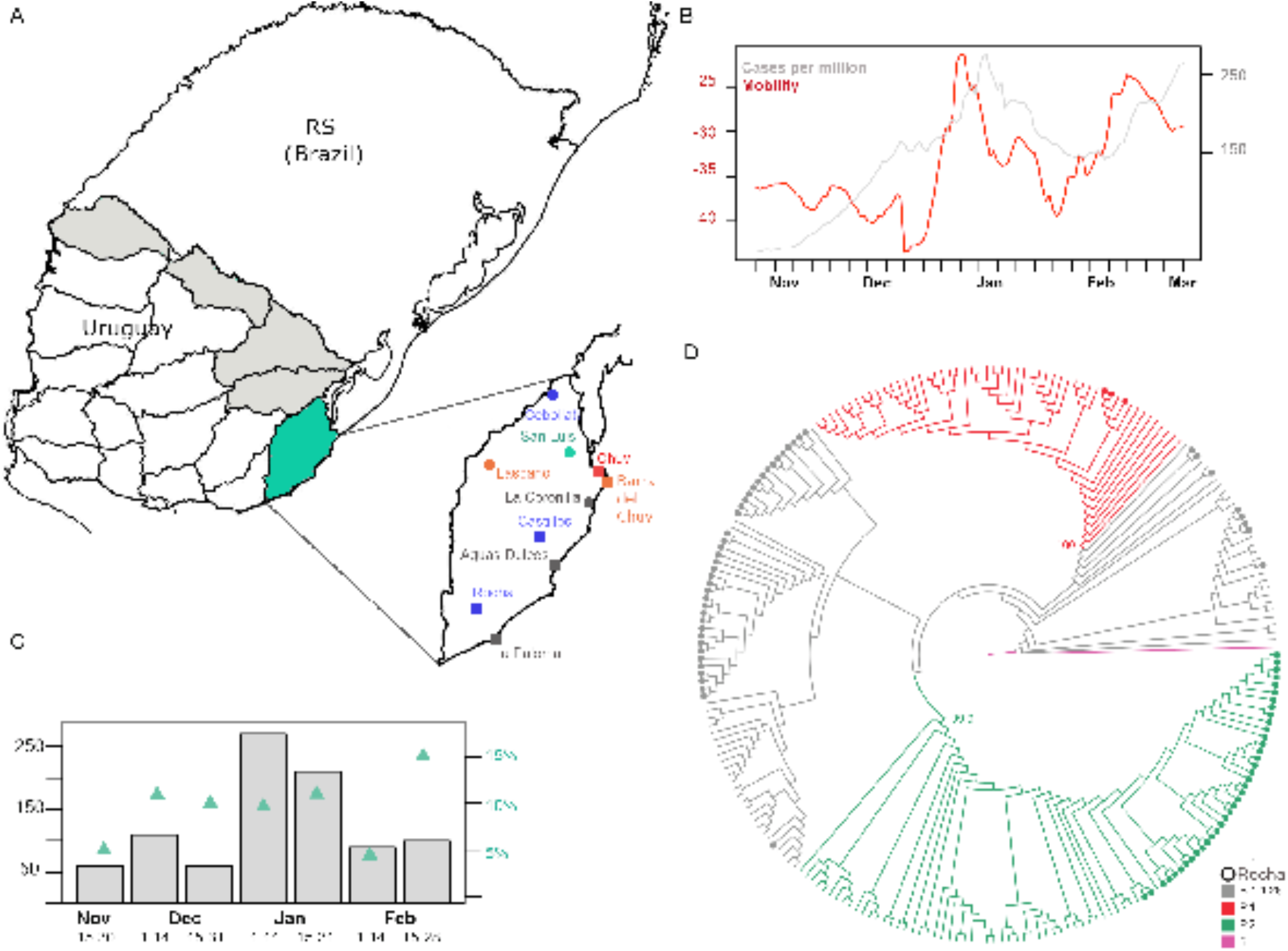
Information on samples and epidemic dynamics of the period of interest. **A**. Map of Uruguay and the Brazilian southern state of Rio Grande do Sul. In gray Uruguayan departments bordering Brazil (dry border). In green, the Uruguayan department of Rocha. To the right, Rocha is zoomed-in and the sample collection sites are marked with dots. Gray dots are the localities with (only) the B.1.1.28 lineage, green dots localities with (only) P.2 variant, blue dots localities with B.1.1.28 and P.2 and orange dots P.1 samples. The red dot corresponds to a location (Chuy) with all three P.1, P.2 and B.1.1.28. Dots shape represents the presence or absence of the novel Q675H and Q677H spike mutation. Square shape represents the presence of the mutations, round shape the absence. **B**. MLtree of lineages B.1.1.28, P.2 and P.1 found in Brazil and Uruguay. Tree was rooted with Wuhan’s reference (blue). B.1.1.28 samples are in gray, P.2 samples in green and P.1 samples in red. The dots in the leaves represent Uruguayan samples. **C**. Barplot of the total number of positive cases per fortnight in Rocha. Green points represent the percentage of cases that were sequenced per fortnight. D. Daily mobility index (red) and number of cases per million (gray) is shown for the time period between 15 November and 28 of February. Mobility index is calculated as in https://hdl.handle.net/20.500.12008/27166.

This study was approved by Ethics Committees in Uruguay (SASA Ethics Committee; supplementary materials).

### SARS-CoV-2 RNA identification from positive samples

Molecular detection of the virus was performed with different kits at each center. In the UdelaR laboratory located in Rocha (CURE Regional Este-Rocha, Uruguay) which is part of the UdelaR Labs Network for SARS-CoV-2 molecular detection, the viral RNA extraction was performed directly from samples (virus transport media containing the swab collected) by using the Viral RNA Isolation Kit (Liferiver Bio-Tech Corp., San Diego, USA), according to the manufacturer’s instructions. The viral RNA extracted was used as a template in a OneStep RT-qPCR kit COVID-19 RT-PCR Real TM Fast (UdelaR, IPMON and ATGen S.R.L., Montevideo, Uruguay) to test the presence of SARS-CoV-2 RNA in the samples, according to the manufacturer’s instructions. Sanatorio Americano Montevideo (SASA, Laboratory of Molecular Biology) used on the one hand, Coronavirus COVID-19 genesig® Real-Time PCR assay and on the other, GeneFinder Covid-19 Plus RealAmp Kit, according to manufacturer’s instructions. The Viral RNA was extracted manually from 140 μl of clinical RT-qPCR positive samples using QIAamp Viral RNA Mini kit (QIAGEN, Hilden, Germany) or automatedly using 300 μl of the sample and Perkin-Elmer Chemagic machine/chemistry, according to the manufacturer’s instructions. Total RNA from positive samples was reverse transcribed using SuperScript™ IV First Strand Synthesis System (Invitrogen, Carlsbad, CA, USA), according to the manufacturer’s instructions.

### SARS-CoV-2 whole-genome amplification

The Viral RNA was extracted manually from 140 μl of clinical RT-qPCR positive samples using QIAamp Viral RNA Mini kit (QIAGEN, Hilden, Germany) or automatedly using 300 μl of the sample and Perkin-Elmer Chemagic machine/chemistry, according to the manufacturer’s instructions. Total RNA from positive samples was reverse transcribed using SuperScript™ IV First Strand Synthesis System (Invitrogen, Carlsbad, CA, USA), according to the manufacturer’s instructions.

Two protocols were used to recover SARS-CoV-2 genomes: long (2kb) and short (400b) PCR amplicon protocols. When SARS-CoV-2 genome amplification was unsuccessful with the long 2kb amplicons strategy, the short amplicon strategy was used.

Long amplicon protocol: Two multiplex PCR reactions with the primer scheme (Pool A = nine amplicons and Pool B = eight amplicons) were performed for building long (∼2kb) amplicon libraries to recover SARS-CoV-2 genomes as previously described by Resende and co-workers [19], by using the Q5® High-Fidelity DNA Polymerase (New England Biolabs, Ipswich, MA, USA), according to the manufacturer’s instructions. Short amplicon protocol: In this case SARS-CoV-2 genomes were recovered by building short (∼400pb) amplicon libraries as previously described by Quick and co-workers [20], using the hCoV-19 primer scheme V3 (https://github.com/artic-network/primer-schemes/tree/master/nCoV-2019/V3). Both protocols (long and short amplicons libraries) are based on the amplicon tiling strategy described previously by Quick and co-workers [21]. Both pools were mixtured and the amplicons were purified using Agencourt AMPure XP beads (Beckman Coulter™, Brea, CA, USA). and a quality control was performed to measure the quantity of DNA using the Qubit™ dsDNA BR Assay Kit (Invitrogen) and Qubit Fluorometric Quantification.

### SARS-CoV-2 whole-genome sequencing

Libraries were prepared by using the Ligation Sequencing Kit (SQK-LSK109) and Native Barcoding Expansion (EXP-NBD104 and EXP-NBD114), both from ONT (Oxford Nanopore Technologies, United Kingdom). The NEBNext Ultra II End Repair/dA-Tailing Module and NEBNext Ultra II Ligation Module were used to ligate barcodes and sequence adapters to each sample (New England Biolabs, Ipswich, MA, USA), according to the manufacturer’s instructions. Negative controls using H2O as template (no RNA) in the RT step were added as amplification controls and were kept in the rest of the process as sequencing controls. We used MinION sequencing platforms (Oxford Nanopore Technologies Ltd., Oxford, UK) as described in detail elsewhere [21]

### SARS-CoV-2 whole-genome consensus sequences

Whole-genome consensus sequences obtained from ONT were generated using an adaptation of the nCoV-2019 novel coronavirus Artic bioinformatics protocol (https://artic.network/ncov-2019/ncov2019-bioinformatics-sop.html) as in [11] and adjustments made by Ferrés in https://github.com/iferres/ncov2019-artic-nf. Positions of interest were manually inspected to resolve undetermined bases. All genomes obtained in this study were uploaded at the EpiCoV database in the GISAID. Accession numbers are in Table S1.

### SARS-CoV-2 genotyping

Uruguayan genome sequences were initially assigned to viral lineages according to [22] using the Pangolin web application (https://pangolin.cog-uk.io) and later confirmed using maximum likelihood (ML) phylogenetic analyses. ML phylogenetic analyses were performed with IQ-TREE version 1.6.12 [23]. Branch support was assessed by the approximate likelihood-ratio test based on a Shimodaira–Hasegawa-like procedure (SH-aLRT) with 1,000 replicates [24].

### Phylogenetic and phylogeographic analysis

Uruguayan P.2 genome sequences (n = 38) were analyzed in the context of additional P.2 sequences from Brazil, downloaded from the EpiCoV database of the GISAID initiative [25]. Downloaded sequences (n = 686, see Supplementary GISAID Acknowledgment P.2) were complete, high quality, with full collection date information and sampled between October 01, 2020, and February 28, 2021. Alignment was performed with MAFFT v7.471 [26]. After careful alignment inspection, six sequences showing very divergent genome regions were excluded, keeping 680 P.2 sequences from Brazil (Table S2). In the case of lineage B.1.1.28, 37 Uruguayan sequences produced in this work and nine sequences previously obtained [11] (Table S1) were aligned with 952 additional B.1.1.28 sequences from Brazil, downloaded from GISAID as before, with sampling date before February 28, 2021 (Supplementary GISAID Acknowledgment B.1.1.28). After alignment inspection, six Brazilian sequences were discarded, retaining a final set of 946 B.1.1.28 sequences from Brazil (Table S3). Finally, as most samples belonged to B.1.1.28 and B.1.1.28-derived lineages (P.1 and P.2) we built a global B.1.1.28 alignment with our sequences (46 B.1.1.28, 38 P.2 and three P.1), the reference Wuhan sequence (NC045512) and the earliest 50 Brazilian samples of each lineage B.1.1.28, P.2 and P.1. In the case of P.1, 169 Brazilian sequences collected before February 28, 2021, were downloaded from GISAID (Supplementary GISAID Acknowledgment P.1), and after alignment and control of high-quality sequences, the earliest 50 sequences were retained and incorporated to the global B.1.1.28 alignment (Table S4).

Maximum likelihood (ML) phylogenetic analysis of final 238 B.1.1.28, P.1 and P.2 sequences was conducted using IQ-TREE version 1.6.12 under the GTR+F+I nucleotide substitution model selected by the built-in ModelFinder option [23]. Branch support was assessed by SH-aLRT with 1,000 replicates. The tree was rooted using Wuhan’s reference. In the case of the dataset of 718 P.2 sequences, the ML tree inference was performed under the GTR+F+R2 nucleotide substitution model and the root established with the sequence EPI_ISL_717925 with collection date 2020-10-09. This tree was time-scaled using TreeTime [27] applying a fixed clock rate of 8 × 10^−4^ substitutions/site/year, based on previous estimates [28] (https://virological.org/t/time-dependence-of-sars-cov-2-substitution-rates/542), a skyline coalescent model with eight grid points and keeping polytomies. The time-scaled tree was then employed for the ancestral character state reconstruction (ACR) of epidemic locations with PastML [29], using the Marginal Posterior Probabilities Approximation (MPPA) method with an F81-like model. The same analysis was performed for the final 992 B.1.1.28 sequences. In this case, the selected nucleotide substitution model was also GTR+F+R2 and the ML tree was rooted with sequence EPI_ISL_416036 with collection date 2020-03-05.

Time-scaled Bayesian phylogeographic analyses were next performed to infer the geographical source and dissemination pattern of Uruguayan P.2 and B.1.1.28 samples and to estimate the time of the most recent common ancestors (T_MRCA_) of the main clades. Phylogenetic trees were estimated in BEAST 1.10 [30] using the GTR+F+I, GTR+F and GTR+F+I nucleotide substitution models for clades UY-I_P2_, UY-I_28_, and UY-II_28_, respectively, the nonparametric Bayesian skyline model as the coalescent tree prior [31], a strict molecular clock model with a uniform substitution rate prior (8^−10^ × 10^−4^ substitutions/site/year) and a reversible discrete phylogeographic model [32] with a continuous-time Markov chain (CTMC) rate reference prior [33]. MCMC chains were run for 50 million generations and convergence (Effective Sample Size > 200) in parameter estimates was assessed using TRACER v1.7 [34]. Maximum clade credibility (MCC) trees were summarized with TreeAnnotator v1.10 [35] and visualized using FigTree v1.4.4 (http://tree.bio.ed.ac.uk/software/figtree). Additional visualizations were implemented in the R environment with treeio and ggtree Bioconductor packages [36].

### Determination of SARS-CoV-2 RS Brazilian lineage prevalence

We downloaded high quality and high coverage sequence data from GISAID from the Brazilian state of RS between September 2020 to February 2021 (Supplementary GISAID Acknowledgment RS) and included additional samples from RS used in http://www.genomahcov.fiocruz.br/presenca-das-linhagens-por-estado/. We obtained 333 sequences (47 in September, 51 in October, 155 in November, 18 in December, 22 in January and 41 February), that were further classified using Pangolin. The proportions of each lineage (B.1.1.33, B.1.1.28, P.1, P.2 and Others B) were calculated monthly. “Others B” include B.1, B.1.1, B.1.91 and B.1.1.332 B-descendant lineages.

### Ct analysis incorporating epidemic dynamics

Cycle threshold (Ct) values from RT-PCR tests were obtained for samples isolated between November, 2020 and February, 2021 in CURE Regional Este-Rocha. A total of 761 positive samples were used for the calculations. We tested whether a difference exists between the Cts values before and after the estimated time of introduction of P.2 VOI using a t-test. Additionally, the data available at https://guiad-covid.github.io/data/estadisticasuy/ was used to visualize the epidemic dynamics during the period of interest. A roll-up average (7 days) on the daily new cases per million inhabitants was calculated and plotted. Based on this, two relevant epidemic phases were characterized: exponential growth and pronounced decrease. The first exponential growth occurred between November 15 and November 30, 2020 and the second one (steeper) from December 24, 2020 to January 9, 2021. The first pronounced decrease in daily new cases was from November 30 to December 24, 2020 and the second one from January 9 to February 4, 2021. On this basis, it was tested whether within the same epidemic dynamic (e.g. exponential growth) the Cts differences observed before and after the introduction of P.2 were statistically significant (t-test). We also determined the mobility index of each period as calculated in https://hdl.handle.net/20.500.12008/27166 based on Google data, and the effective reproductive number (Re) for Rocha (7 day roll-up average) as calculated by the EpiEstim model [37].

## RESULTS

### Prevalent SARS-CoV-2 lineages in the southeast Uruguayan-Brazilian border

To understand the dynamics of SARS-CoV-2 spread during the first pandemic wave in Uruguay by the end of November, 2020, when the TETRIS strategy was lost, we sequenced the viral genome from 84 patients diagnosed between 25th November, 2020 and 26th February, 2021, in Rocha department, a sentinel location at the southeast border with Brazil (**Fig 1A** and Table S1). **Fig 1B** and **Fig S1A** depict several attributes of the COVID-19 epidemics in Rocha, such as cases per million inhabitants, mobility index and estimated Re. These sequences represent ∼10% of all laboratory-confirmed cases (n=900) in Rocha department at the studied period and were sampled in a temporally comprehensive manner (**Fig 1C**). Only two of the sequenced samples belong to individuals who reported travel to Brazil, indicating that most of them were locally infected. More than half of the patients (n=47) live in Chuy, a twin Brazilian-Uruguayan city. The SARS-CoV-2 genotyping of Rocha samples revealed that all sequences belonged to the B.1 lineage characterized by the D614G mutation at the S protein and most of them to B.1.1.28 and descendants’ lineages (**Fig 1D** and Table S1). Of note, by the second half of February, 2021, three samples of the lineage P.1 were also detected (**Fig 1D**), representing the first detection of this variant of concern (VOC) in Uruguay up to date [14].

Lineages B.1.1.28 and P.2 were the most prevalent in Rocha, representing 44% and 45% of the samples, respectively (**Fig 2A** and Table S1). Lineage B.1.1.28 was the most prevalent in the period November-December, 2020 while P.2 became the dominant one during January-February, 2021, thus supporting a similar lineage replacement process as observed in several Brazilian states including RS, the southernmost Brazilian state (**Fig 2A)**. As it is highlighted in **Fig 2A**, there is an empirically observed time lag of about three months between P.2 reported in RS and Rocha, which likely depends not only on the variant’s dissemination dynamic but also on the different sequencing efforts. Given the change in the dominant viral variant and to assess whether introduction of the VOI P.2 could have an impact on patients’ viral loads, we compared 761 Ct values within two periods, prior and after first detection of the VOI (Fig S1). We found a statistically significant decrease in Ct values in the after-P.2 period respect to the before-P.2 period (Fig S1B, left panel, p-value <0.0035) and this effect remains when comparison was restricted to phases with equivalent epidemiological scenarios (exponential growth phase, p-value<0.0022) as suggested in [38] (Fig S1B, right panel).

**Figure 2.**
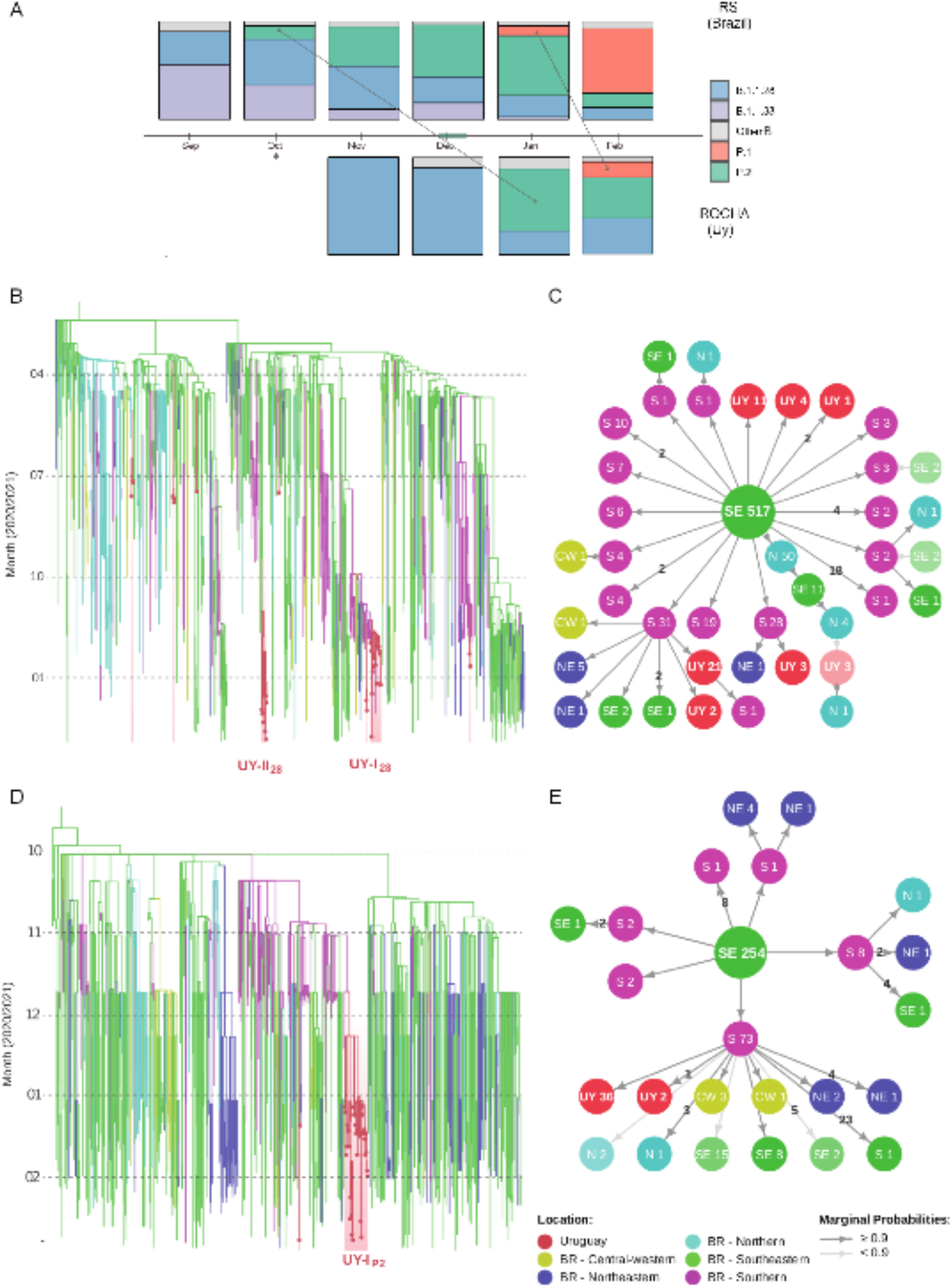
Prevalences and origins of major SARS-CoV-2 lineages in Rocha. **A**. Prevalences of different lineages/variants in RS (Brazil) and Rocha (Uruguay). On top, the monthly variant’s prevalence in RS (starting in September, 2020). On the bottom, the monthly variant’s prevalence in Rocha (starting in November, 2020 since in September there were only 10 cases in Rocha and zero in October, 2020, marked with an asterisk). A first line connects the presence of P.2 in RS with its presence in Rocha (three months lag). A second line connects the detection of P.1 in RS with its detection in Rocha (one month lag). On the time-line the P.2 TMRCA confidence interval is shown as calculated in this study. **B and C**. Origin of major SARS-CoV-2 lineages circulating in Rocha. Maximum likelihood phylogeographic analysis of lineage B.1.1.28 (n=992) inferred by ancestral character reconstruction method implemented in PastML. **B**. Tips and branches are colored according to sampling location and the most probable location state of their descendent nodes, respectively, as indicated in the legend. Shaded boxes highlight the major B.1.1.28 clades in Rocha. The time scaled tree was rooted with the earliest sequence. **C**. Schematic representation of migration events. Each node in the network is identified by location and number of sequences within different phylogenetic subclusters. Arrows indicate migration events deduced from location state changes across the tree. The shade of gray in the arrows identifies marginal probabilities and the numbers quantify the migration events connecting respective locations (no number represents one single event). Nodes are colored according to their location. BR: Brazil. **D and E**. Same as B and C, but the maximum likelihood phylogeographic analysis was performed on sequences (n=718) of VOI P.2. Shaded red box highlights the major P.2 clade in Rocha.

### Identification of the main B.1.1.28 and P.2 clades disseminated in Rocha and their Brazilian origin

A total of 46 B.1.1.28 and 38 P.2 Uruguayan sequences were combined with 946 B.1.1.28 and 680 P.2 Brazilian sequences available in the GISAID database respectively. The ML tree of the B.1.1.28 data set supports at least seven independent introductions of this lineage from Brazil into Uruguay (**Figs 2B and C**). Three introductions resulted in singletons or dyads with no evidence of further local dissemination. Four introductions, however, resulted in highly supported clades (SH-aLRT ≥ 0.85) of 3 to 21 Uruguayan sequences sampled at early 2020 or late 2020/early 2021. None Uruguayan B.1.1.28 clade comprises sequences sampled at both periods. According to the ML-based ACR of epidemic locations, the viral introductions that gave rise to the largest B.1.1.28 characterized clades designated UY-I_28_ (n = 22) and UY-II_28_ (n = 11), were most likely from the Southern and Southeastern Brazilian regions respectively (ACR location marginal probability ≥ 0.9). Clade UY-I_28_ comprises sequences from Castillos (CAS), Cebollatí (CEB), Chuy (CHY), La Coronilla (COR) and Rocha capital city (ROC) from samples isolated between November 25 and February 23. Clade UY-II_28_ is made up of sequences from CAS, CHY, ROC, Aguas Dulces (ADU), Barra del Chuy (BCY) and La Paloma (PAL) from samples isolated between January 12 and February 25.

The ML phylogeographic analysis of the P.2 dataset revealed two independent viral introductions into Uruguay most likely from the Brazilian Southern region (ACR location marginal probability ≥ 0.9) that originated two highly supported Uruguayan P.2 clades (SH-aLRT > 0.85) (**Fig1D** and **Figs 2D and E**). The main clade designated as UY-I_P2_ (n=36) comprises sequences corresponding to samples isolated in CAS, CEB, CHY, COR, ROC and San Luis al Medio (SLU), between January 5 and February 24, 2021, while the minor clade designated UY-II_P2_ (n=2), comprises sequences sampled at Rocha and Rivera Uruguayan departments.

### Introduction and transmission of lineage B.1.1.28 in Rocha in November 2020

Bayesian phylogeographic analyses of the main B.1.1.28 clades detected in Rocha (UY-I_28_ and UY-II_28_) along with its basal Brazilian sequences according to the ML inference are shown in **Figs 3A and B**. The analysis suggests that clade UY-I_28_ arose after the introduction of a viral strain from the Southern region of Brazil to the Uruguayan city of Chuy (Posterior State Probability [PSP] = 0.96) around November 23, 2020 (95% Highest Probability Density [HPD]: November 12 to November 23, 2020), that subsequently spread to the rest of the localities of Rocha (**Fig 3A**). Only one synonymous mutation (Orf1ab:C8683T) distinguishes UY-I_28_ Rocha sequences from its basal ones corresponding to RS samples.Sequences in this clade also present the P13L change in the Nucleocapsid viral protein reported previously as widely present in B.1.1.28 sequences from RS [16,17]. The spatiotemporal reconstruction of UY-II_28_ suggests that this clade probably arose around November 3, 2020 (HPD: September 24 to December 6, 2020) with a viral introduction into the city of Rocha (capital of the homonymous department) from the southeastern Brazilian region (PSP = 0.79) (**Fig 3B**). Notably, this sub-clade shows many clade-defining mutations, including two non-synonymous changes in the S protein (Q675H and Q677H).

**Figure 3.**
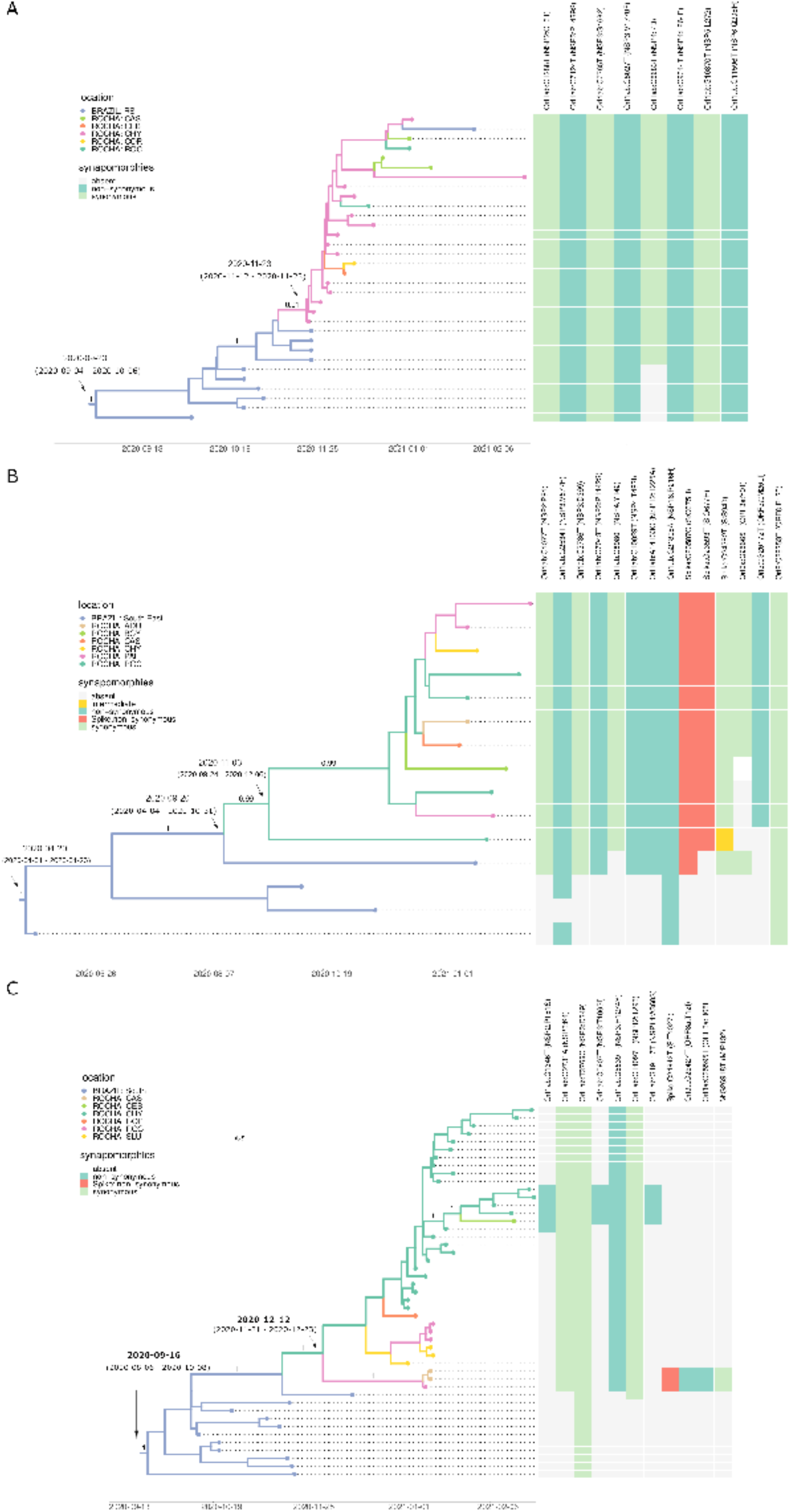
Phylogeographic analyses. Bayesian phylogeographic analysis of main P.2 and B.1.1.28 clades found in Rocha, implemented in BEAST. **A**. Uruguayan P.2 sequences in clade UY-IP2 were combined with 11 additional basal sequences from Southern Brazil. Tips and branches of the time-scales Bayesian tree are colored according to sampling location and the most probable location state of their descendent nodes, respectively, as indicated in the legend. Posterior probability support values and estimated TMRCAs are indicated at key nodes. Additionally, a heatmap represents the presence or absence of synapomorphic sites. The color scheme indicates the different mutations, as indicated in the legend. In each case, genomic position, nucleotide substitution, viral protein and amino acid is shown. **B**. Same as A, but for Uruguayan B.1.1.28 sequences in clade UY-I28 which were combined with 11 additional basal sequences from RS. **C**. Same as A, but for Uruguayan B.1.1.28 sequences in cladeUY-II28 which were analyzed together with four additional basal sequences from South Eastern Brazil. In the heatmap, the yellow square highlights a sample where in genomic position 24,382, both reference C and alternative T, were equally probable.

### Introduction and transmission of lineage P.2 in Rocha in December 2020

The time scaled phylogeographic analysis of the UY-I_P2_ clade and its basal Brasilian sequences traced its origin to December 12, 2020 (95% HPD: November 21 to December 23, 2020) and Chuy is suggested as the most likely location for the viral introduction (PSP = 0.96) from the Brazilian Southern region (**Fig 3C**). Synapomorphies defining the Rocha UY-I_P2_ clade are three synonymous mutations in Orf1ab and one non-synonymous mutation in Orf1ab (NSP3:H1274Y). Of note, a sub-cluster of sequences presents an additional non-synonymous mutation in the S protein S:T1027I already described as one of the 21 lineage-defining mutations of VOC P.1 (**Fig 3C**) [4,5].

## DISCUSSION

In this work we have analyzed the SARS-CoV-2 viral dissemination between November 2020 and February 2021 in Rocha, a sentinel department in Eastern Uruguay that can be considered as one of the viral getaways from Southern Brazil to the country. By comparing lineage prevalence in the Southernmost Brazilian state of RS and Rocha, we observed a similar molecular epidemiological pattern between both locations with a temporal delay (e.g., three months for P.2 and one month for P.1). The VOI P.2 was first detected in RS in October 2020 at low prevalence (14%) and became the most frequent variant in December 2020 (53%). In January, 2021, the introduction of P.1 was established and it rapidly dominated over other variants in RS from February (66%) onwards. In Rocha, we identified a first period with dominance of lineage B.1.1.28 in November-December, 2020 and a second period with dominance of P.2 in January-February, 2021, while the first P.1 sequences were detected in February, 2021 [14]. The observed time lag would vary depending on the epidemiological genomic surveillance effort in each country and also on the dissemination pattern and biological factors associated with each viral lineage and specific variant, as can be seen for P.1 and P.2 cases.

It has been suggested that after the introduction of border restrictions to human mobility in Uruguay, the viruses on each side of the border separated into different genetic clusters [16]. Our study, however, demonstrates a frequent viral flux between Rocha and the RS during the studied period. The shared epidemic pattern herein detected between Southern Brazil and Rocha is probably extended to other Uruguayan departments bordering Brazil, as we found in our previous study conducted at the departments of Rivera, Artigas and Treinta y Tres [11]. This pattern may not be necessarily valid throughout the whole country as, for instance, in Montevideo, the country’s capital city, the presence of an international airport might have fueled the introduction and dissemination of SARS-CoV-2 strains from further abroad [10,39]. While this work does not assess the SARS-CoV-2 variants circulating through the whole country and additional studies are needed, our study supports that Brazil will stay the most important source of viral diversity in Uruguay.

Towards the end of 2020, a remarkable exponential growth in COVID-19 daily cases was observed, being considered as the primary wave for Uruguay, with the consequent loss of the TETRIS safety zone. Our estimates suggest that in Rocha, this epidemiological worsening was likely associated with the introduction and dissemination of two B.1.1.28 clades. The main clade was probably introduced from RS through Chuy, a Uruguayan-Brazilian twin-city, around late November 2020; while the second clade was probably introduced in the capital city of Rocha around early November 2020. The major B.1.1.28 clade detected in Rocha (UY-I_28_) displayed only one lineage-defining synonymous mutation (Orf1ab:C8683T) in addition to a N:P13L change that is widely present in B.1.1.28 sequences from RS [16, 17], features that are consistent with a probable route of introduction from RS. The minor B.1.1.28 cluster (UY-II_28_), in contrast, was distinguished by several mutations, including two amino acid changes in the S protein: Q675H and Q677H. Mutation S:Q675H was detected in a basal sequence from São Paulo state, but mutation S:Q677H was exclusive of Uruguayan B.1.1.28 sequences.

The amino acid change S:Q677H has been reported as a recurrent mutation arising independently in many SARS-CoV-2 lineages circulating worldwide by the end of 2020, (https://virological.org/t/detection-of-the-recurrent-substitution-q677h-in-the-spike-protein-of-sars-cov-2-in-cases-descended-from-the-lineage-b-1-429/660, [40]). The convergent evolution in different viral lineages and the proximity to the polybasic cleavage site at the S1/S2 boundary raise concern about the potential functional relevance and impact on viral spread of that mutation. We are not aware of previous reports of mutation S:Q675H nor both mutations (Q675H and Q677H) together in emergent SARS-CoV-2 VOI or VOCs. A recent study, however, that developed a model on epidemiological variables which integrates the effects of S amino acid changes in viral fitness, predicted that both S:Q675H and S:Q677H mutations could appear in emerging SARS-CoV-2 VOCs in the following months [41]. Although our phylogeographic analysis inferred the introduction of clade UY-II_28_ from Southeast Brazil into the Rocha department’s capital, we could not rule out the existence of intermediate viral migration steps that were not recovered because of sampling gaps. Given the unlikely existence of a direct link between southeastern Brazil and Rocha, a prior dissemination of this sub-lineage in other parts of Uruguay before being introduced in Rocha seems like a reasonable assumption. Indeed, this might have contributed to the emergence of the Uruguayan first COVID-19 wave by the end of 2020.

By January, 2021, we confirmed the circulation of VOI P.2 in Rocha. This variant harbors the E484K amino acid change in the receptor-binding domain (RBD) of the viral Spike among its five lineage-defining mutations [6]. S:E484K has emerged independently in multiple VOCs (P.1, B.1.351 and B.1.1.7) and VOIs (P.2 and B.1.526) and has been identified as one of the mutations of greatest concern, since it might confer a fitness advantage to the virus by contributing to its immune evasion and increasing its transmissibility by enhanced affinity of RBD to human ACE2 receptor [42-48]. Very soon after its description, evidence about P.2 being involved in reinfection cases was available [49,50]. In this work, all P.2 sequences from Rocha, but one, belonged to a major clade introduced from Southern Brazil through the Chuy city around December 12, 2020, which concurs with P.2 already being the main viral variant in Rocha in January, 2021. These findings confirm the relevance of the bordering city of Chuy as a key sentinel location for early detection of new SARS-CoV-2 variants introduced in Uruguay.

We found a sharp increase of P.2 frequency in Rocha, with partial replacement of earlier circulating B.1.1.28 lineages, as previously reported in RS and many other Brazilian states [7,15-17]. The dissemination of the VOI P.2 in Rocha may have been facilitated by the notorious increment in mobility and lax social distancing in that department (a popular holiday destination for Uruguayans) that occurred at the beginning of 2021. Furthermore, the VOI P.2 may have an increased transmissibility as suggested by the observed drop in Ct values in our dataset; although this finding should be interpreted with caution due to several potential confounding factors (as patient sex and age and variable times from symptoms onset to sample collection) that could not be assessed with our available metadata. The combination of more transmissible variants in a context of high mobility has already been suggested as a key element underlying COVID-19 waves, as proposed for the successful P.1 dissemination in the Amazonas state and in Uruguay [5,14].

Even though Uruguayan borders were closed for tourism, this work shows that SARS-CoV-2 lineages circulating in Rocha, a department in Eastern Uruguay, were representative of what was circulating in RS, Southern Brazil; which concurs with what we found in our previous work in other border departments. We see about a one to two months delay between the arrival of the emergent P.1 and P.2 lineages in RS and its subsequent detection in Rocha. We further demonstrate that the VOI P.2 and one B.1.1.28 clade was in fact introduced through the bi-national city of Chuy, where close economic, social and cultural contact with the Brazilian side is prone to enable recurrent viral flux between both countries. These findings provide clear evidence that a Uruguayan genomic surveillance program should be undertaken in a systematic fashion, specifically allocating resources on these key border locations, to detect new viral lineage introductions in real-time, thus allowing the rapid implementation of measures to achieve long-term COVID-19 epidemic control in Uruguay.

## Data Availability

All genome sequences obtained in this study were uploaded at the EpiCoV database in the GISAID initiative. All genomes obtained in this study were uploaded at the EpiCoV database in the GISAID. Accession numbers are in Table S1.

## Acknowledgment

Thanks for the efforts of the groups that contributed SARS-CoV-2 genomes to the EpiCoV GISAID initiative (Supplementary GISAID Acknowledgement Tables S5-S8).

We thank Ignacio Ferrés for bioinformatic pipeline sharing, Ernesto Mordecki for helpful statistical analysis and Marcelo Fiori for data sharing and helpful discussions.

We specially thank Matias Castells, Matias Salvo and Andres Lizasoain for sample collection and discussion.

## Ethics statement

All relevant ethical guidelines have been appropriately followed. The project was approved by the Ethics Committee of the Sanatorio Americano (Uruguay), ethical approval was given and signed informed consent was obtained from the participants.

## References

[1] Darlan S. Candido, Ingra M. Claro, Jaqueline G. De Jesus, William M. Souza, Filipe R. R. Moreira, et al. Evolution and epidemic spread of SARS-CoV-2 in Brazil. Science 2020: 1255–1260

[2] Paola Cristina Resende, Edson Delatorre, Tiago Gräf, Daiana Mir, Fernando Couto Motta, et al. Evolutionary Dynamics and Dissemination Pattern of the SARS-CoV-2 Lineage B.1.1.33 During the Early Pandemic Phase in Brazil. Frontiers in Microbiology 2021, 2021:3565

[3] T Fujino, H Nomoto, S Kutsuna, M Ujiie, T Suzuki, et al. Novel SARS-CoV-2 Variant in Travelers from Brazil to Japan. Emerg Infect Dis. 2021 Apr;27(4):1243–1245. doi: 10.3201/eid2704.210138.

[4] Nuno R Faria, Thomas A Mellan, Charles Whittaker, Ingra M Claro, Darlan da S Candido, et al. Genomics and epidemiology of the P.1 SARS-CoV-2 lineage in Manaus, Brazil. Science 2021, 2021:815--821. doi: 10.1126/science.abh2644.

[5] FG Naveca, V Nascimento, VC de Souza, AL Corado, F Nascimento, et al. COVID-19 in Amazonas, Brazil, was driven by the persistence of endemic lineages and P.1 emergence. Nat Med. 2021. doi: 10.1038/s41591-021-01378-7.

[6] CM Voloch, FR Jr da Silva, LGP de Almeida, CC Cardoso, OJ Brustolini, et al. Genomic characterization of a novel SARS-CoV-2 lineage from Rio de Janeiro, Brazil. J Virol. 2021, 95(10):e00119–21. doi: 10.1128/JVI.00119-21.

[7] AP Lamarca, LGP de Almeida, Ronaldo da Silva Francisco, LFA Lima, KC Scortecci, et al. Genomic surveillance of SARS-CoV-2 tracks early interstate transmission of P.1 lineage and diversification within P.2 clade in Brazil. medRxiv 2021. doi:10.1101/2021.03.21.21253418

[8] VB Franceschi, PAG Ferrareze, RA Zimerman, GB Cybis, CE Thompson. Mutation hotspots, geographical and temporal distribution of SARS-CoV-2 lineages in Brazil, February 2020 to February 2021: insights and limitations from uneven sequencing efforts. medRxiv 2021. doi:10.1101/2021.03.08.21253152

[9] Luke Taylor. Why Uruguay lost control of COVID. Nature 2021, 2021: 21. doi: https://doi.org/10.1038/d41586-021-01714-4

[10] V Elizondo, GW Harkins, B Mabvakure, S Smidt, P Zappile, et al. SARS-CoV-2 genomic characterization and clinical manifestation of the COVID-19 outbreak in Uruguay. Emerg Microbes Infect. 2021, 10(1):51–65. doi: 10.1080/22221751.2020.1863747.

[11] D Mir D, N Rego, PC Resende, F Tort, T Fernández-Calero, et al. Recurrent Dissemination of SARS-CoV-2 Through the Uruguayan-Brazilian Border. Front Microbiol. 2021 May 28;12:653986. doi: 10.3389/fmicb.2021.653986.

[12] C Fraser, S Riley, RM Anderson, NM Ferguson. Factors that make an infectious disease outbreak controllable. Proc Natl Acad Sci U S A. 2004, 101(16):6146–51. doi: 10.1073/pnas.0307506101.

[13] KH Grantz, EC Lee, M. D’Agostino, KH Lee, CJE Metcalf, et al. Maximizing and evaluating the impact of test-trace-isolate programs: A modeling study. PLoS Med. 2021, 18(4):e1003585. doi: 10.1371/journal.pmed.1003585.

[14] N Rego, A Costábile, M Paz, C Salazar, P Perbolianachis, et al. Implementation of a qPCR assay coupled with genomic surveillance for real-time monitoring of SARS-CoV-2 variants of concern. medRxiv 2021. doi: 10.1101/2021.05.20.21256969

[15] VB Franceschi, GD Caldana, A de Menezes Mayer, GB Cybis, CAM Neves, et al. Genomic epidemiology of SARS-CoV-2 in Esteio, Rio Grande do Sul, Brazil. BMC Genomics. 2021 May 20;22(1):371. doi: 10.1186/s12864-021-07708-w.

[16] RDS Jr Francisco, LF Benites, AP Lamarca, LGP de Almeida, AW Hansen, et al. Pervasive transmission of E484K and emergence of VUI-NP13L with evidence of SARS-CoV-2 co-infection events by two different lineages in Rio Grande do Sul, Brazil. Virus Res. 2021 Apr 15;296:198345. doi: 10.1016/j.virusres.2021.198345.

[17] F. Sant’Anna, AP Muterle Varela, J Prichula, J Comerlato, C Comerlato, et al. Emergence of the novel SARS-CoV-2 lineage VUI-NP13L and massive spread of P.2 in South Brazil. Emerging Microbes & Infections, 2021, doi: 10.1080/22221751.2021.1949948.

[18] VB Franceschi, GD Caldana, C Perin, A Horn, C Peter, et al. Predominance of the SARS-CoV-2 lineage P.1 and its sublineage P.1.2 in patients from the metropolitan region of Porto Alegre, Southern Brazil in March 2021: a phylogenomic analysis. medRxiv 2021, doi: 10.1101/2021.05.18.21257420.

[19] PC Resende. Long reads nanopore sequencing to recover SARS-CoV-2 whole genome V.3. BioRxiv 2020, dx.doi.org/10.17504/protocols.io.bfy7jpzn

[20] Josh Quick. nCoV-2019 sequencing protocol v2 (GunIt) V.2. protocols.io 2020. dx.doi.org/10.17504/protocols.io.bdp7i5rn

[21] J Quick, ND Grubaugh, S. Pullan, IM Claro, AD Smith, et al. Multiplex PCR method for MinION and Illumina sequencing of Zika and other virus genomes directly from clinical samples. Nat Protoc. 2017 Jun;12(6):1261–1276. doi: 10.1038/nprot.2017.066.

[22] A Rambaut, EC Holmes, A O’Toole, V Hill, JT McCrone, et al. A dynamic nomenclature proposal for SARS-CoV-2 lineages to assist genomic epidemiology. Nat Microbiol. 2020 Nov;5(11):1403–1407. doi: 10.1038/s41564-020-0770-5. Epub 2020 Jul 15. PMID: 32669681; PMCID: PMC7610519.

[23] LT Nguyen, HA Schmidt, A von Haeseler, BQ Minh. IQ-TREE: a fast and effective stochastic algorithm for estimating maximum-likelihood phylogenies. Mol Biol Evol. 2015 Jan;32(1):268–74. doi: 10.1093/molbev/msu300. Epub 2014 Nov 3. PMID: 25371430; PMCID: PMC4271533.

[24] M Anisimova, O Gascuel. Approximate likelihood-ratio test for branches: A fast, accurate, and powerful alternative. Syst Biol. 2006 Aug;55(4):539–52. doi: 10.1080/10635150600755453. PMID: 16785212.

[25] Y Shu, J McCauley. GISAID: Global initiative on sharing all influenza data - from vision to reality. Euro Surveill. 2017 Mar 30;22(13):30494. doi: 10.2807/1560-7917.ES.2017.22.13.30494.

[26] K Katoh, DM Standley. MAFFT multiple sequence alignment software version 7: improvements in performance and usability. Mol Biol Evol. 2013 Apr;30(4):772–80. doi: 10.1093/molbev/mst010. Epub 2013 Jan 16. PMID: 23329690; PMCID: PMC3603318.

[27] P Sagulenko, V Puller, RA Neher. TreeTime: Maximum-likelihood phylodynamic analysis. Virus Evol. 2018 Jan 8;4(1):vex042. doi: 10.1093/ve/vex042. PMID: 29340210; PMCID: PMC5758920.

[28] S Duchene, L Featherstone, M Haritopoulou-Sinanidou, A Rambaut, P Lemey, et al. Temporal signal and the phylodynamic threshold of SARS-CoV-2. Virus Evol. 2020 Aug 19;6(2):veaa061. doi: 10.1093/ve/veaa061. PMID: 33235813; PMCID: PMC7454936.

[29] SA Ishikawa, A Zhukova, W Iwasaki, O Gascuel. A Fast Likelihood Method to Reconstruct and Visualize Ancestral Scenarios. Mol Biol Evol. 2019 Sep 1;36(9):2069–2085. doi: 10.1093/molbev/msz131. PMID: 31127303; PMCID: PMC6735705.

[30] MA Suchard, P Lemey, G Baele, DL Ayres, AJ Drummond et al. Bayesian phylogenetic and phylodynamic data integration using BEAST 1.10. Virus Evol. 2018 Jun 8;4(1):vey016. doi: 10.1093/ve/vey016. PMID: 29942656; PMCID: PMC6007674.

[31] AJ Drummond, A Rambaut, B Shapiro, OG Pybus. Bayesian coalescent inference of past population dynamics from molecular sequences. Mol Biol Evol. 2005 May;22(5):1185–92. doi: 10.1093/molbev/msi103. Epub 2005 Feb 9. PMID: 15703244.

[32] P Lemey, A Rambaut, AJ Drummond, MA Suchard. Bayesian phylogeography finds its roots. PLoS Comput Biol. 2009 Sep;5(9):e1000520. doi: 10.1371/journal.pcbi.1000520. Epub 2009 Sep 25. PMID: 19779555; PMCID: PMC2740835.

[33] MAR Ferreira, MA Suchard. Bayesian analysis of elapsed times in continuous-time Markov chains. The Canadian Journal of Statistics2010. doi: 10.1002/cjs.5550360302

[34] A Rambaut, AJ Drummond, D Xie, G Baele, MA Suchard. Posterior Summarization in Bayesian Phylogenetics Using Tracer 1.7. Syst Biol. 2018 Sep 1;67(5):901–904. doi: 10.1093/sysbio/syy032.

[35] R Bouckaert, TG Vaughan, J Barido-Sottani, S Duchêne, M Fourment, et al. BEAST 2.5: An advanced software platform for Bayesian evolutionary analysis. PLoS Comput Biol. 2019 Apr 8;15(4):e1006650. doi: 10.1371/journal.pcbi.1006650.

[36] G Yu, DK Smith, H Zhu, Y Guan, TT Lam. ggtree: an r package for visualization and annotation of phylogenetic trees with their covariates and other associated data. Methods in Ecology and Evolution2016. doi: 10.1111/2041-210X.12628

[37] RN Thompson, JE Stockwin, RD van Gaalen, JA Polonsky, ZN Kamvar. Improved inference of time-varying reproduction numbers during infectious disease outbreaks. Epidemics. 2019 Dec;29:100356. doi: 10.1016/j.epidem.2019.100356.

[38] JA Hay, L Kennedy-Shaffer, S Kanjilal,NJ Lennon, SB Gabriel, et al. Estimating epidemiologic dynamics from cross-sectional viral load distributions. Science 2021. doi: 10.1126/science.abh0635

[39] C Salazar, A Costabile, I Ferrés, P Perbolianachis, M Pereira-Gómez, et al. Case Report: Early Transcontinental Import of SARS-CoV-2 Variant of Concern 202012/01 (B.1.1.7) From Europe to Uruguay. Frontiers in Virology 2021, doi: 10.3389/fviro.2021.685618

[40] MR Farcet, M Karbiener, J Schwaiger, R Ilk, TR Kreil, Thomas R. Rapidly Increasing SARS-CoV-2 Neutralization by Intravenous Immunoglobulins Produced from Plasma Collected During the 2020 Pandemic. bioRxiv 2021, doi: 10.1101/2021.02.12.430933

[41] MC Maher, I Bartha, S Weaver, J di Iulio, E Ferri, et al. Predicting the mutational drivers of future SARS-CoV-2 variants of concern. medRxiv 2021, doi: 10.1101/2021.06.21.21259286

[42] A Baum, BO Fulton, E Wloga, R Copin, KE Pascal, et al. Antibody cocktail to SARS-CoV-2 spike protein prevents rapid mutational escape seen with individual antibodies. Science. 2020 Aug 21;369(6506):1014–1018. doi: 10.1126/science.abd0831.

[43] S Jangra, C Ye, R Rathnasinghe, D Stadlbauer, Personalized Virology Initiative study group, et al. SARS-CoV-2 spike E484K mutation reduces antibody neutralisation. Lancet Microbe. 2021 Apr 7. doi: 10.1016/S2666-5247(21)00068-9.

[44] AJ Greaney, TN Starr, P Gilchuk, SJ Zost, E Binshtein, et al. Complete Mapping of Mutations to the SARS-CoV-2 Spike Receptor-Binding Domain that Escape Antibody Recognition. Cell Host Microbe. 2021 Jan 13;29(1):44-57.e9. doi: 10.1016/j.chom.2020.11.007.

[45] H Gu, Q Chen, G Yang, L He, H Fan, et al. Adaptation of SARS-CoV-2 in BALB/c mice for testing vaccine efficacy. Science. 2020 Sep 25;369(6511):1603–1607. doi: 10.1126/science.abc4730.

[46] K Leung, MH Shum, GM Leung, TT Lam, JT Wu. Early transmissibility assessment of the N501Y mutant strains of SARS-CoV-2 in the United Kingdom, October to November 2020. Euro Surveill. 2021 Jan;26(1):2002106. doi: 10.2807/1560-7917.ES.2020.26.1.2002106.

[47] G Nelson, O Buzko, P Spilman, K Niazi, S Rabizadeh, et al. Molecular dynamic simulation reveals E484K mutation enhances spike RBD-ACE2 affinity and the combination of E484K, K417N and N501Y mutations (501Y.V2 variant) induces conformational change greater than N501Y mutant alone, potentially resulting in an escaellipsis. bioRvix 2021, 10.1101/2021.01.13.426558

[48] PAG Ferrareze, VB Franceschi, AM Mayer, GD Caldana, RA Zimerman, et al. E484K as an innovative phylogenetic event for viral evolution: Genomic analysis of the E484K spike mutation in SARS-CoV-2 lineages from Brazil. Infect Genet Evol. 2021 May 25;93:104941. doi: 10.1016/j.meegid.2021.104941.

[49] CKV Nonaka, MM Franco, T Gräf, CA de Lorenzo Barcia, RN de Ávila Mendonça, et al. Genomic Evidence of SARS-CoV-2 Reinfection Involving E484K Spike Mutation, Brazil. Emerg Infect Dis. 2021 May;27(5):1522–1524. doi: 10.3201/eid2705.210191.

[50] PC Resende, JF Bezerra, RH Teixeira Vasconcelos, I Arantes, L Appolinario, et al. Severe Acute Respiratory Syndrome Coronavirus 2 P.2 Lineage Associated with Reinfection Case, Brazil, June-October 2020. Emerg Infect Dis. 2021 Jul;27(7):1789–1794. doi: 10.3201/eid2707.210401.

