## Supplementary material for "Spatiotemporal dissemination pattern of SARS-CoV-2 B1.1.28-derived lineages introduced into Uruguay across its southeastern border with Brazil": fig S1

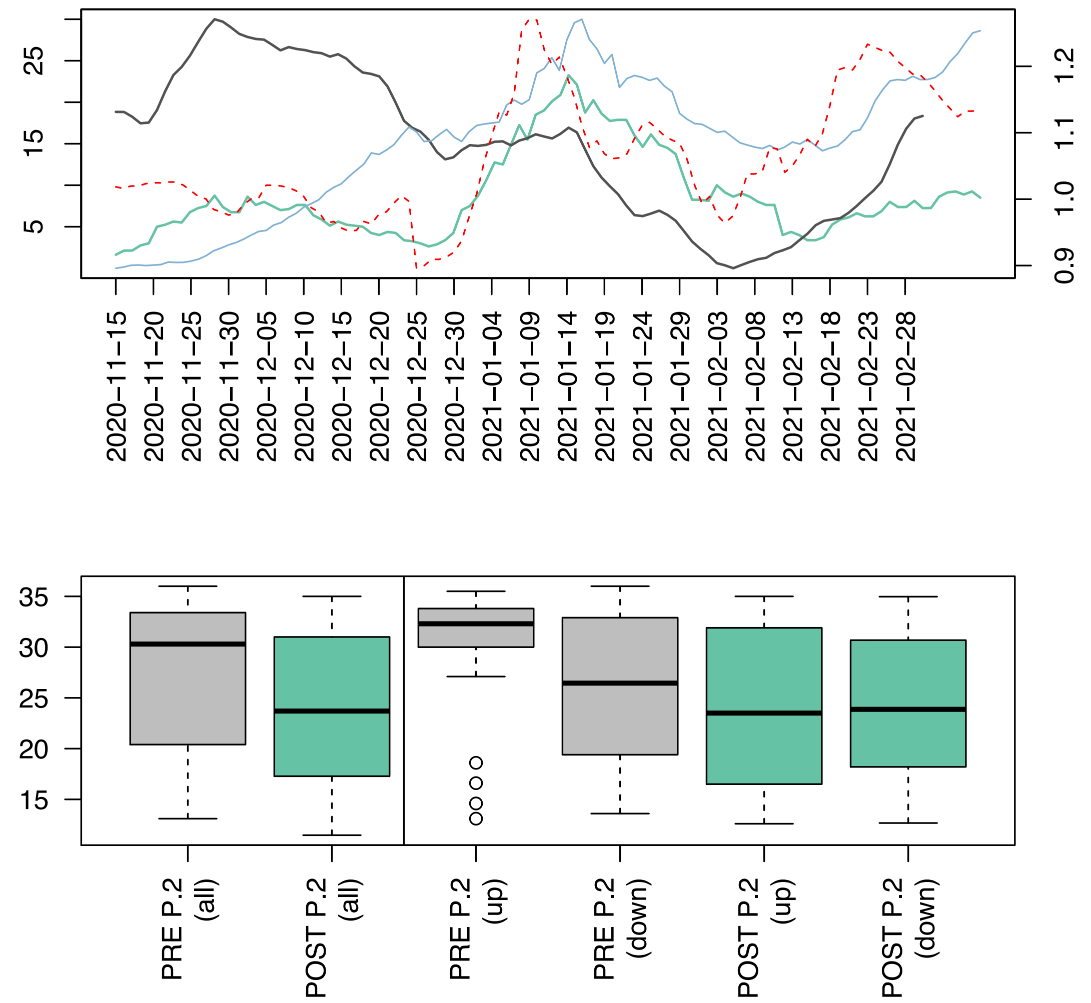


**Supplementary Figure S1**. **Ct values in different epidemic phases A.** In black effective reproductive number (Re) estimations for Rocha as calculated by EpiEstim model for the time period between 2020-11-15 and 2021-02-28. In red, the mobility index for Rocha as calculated by GUIAD group (https://hdl.handle.net/20.500.12008/27166) for the same period. Green and blue lines represent new daily cases per million (7-day rollup average) for Rocha and Uruguay, respectively. **B.** The Cycle threshold (Ct) values for 761 positive samples isolated between November, 2020 and February, 2021 in CURE

Regional Este-Rocha are considered. Boxplots showing Cts values distributions before and after the estimated time of introduction of VOI P.2 are shown in gray (before) and green (after). The left panel includes all samples before and after VOI P.2 introduction. Right panel shows the distributions for the previously characterized epidemic phases (up and down stands for exponential growth and pronounced decrease, respectively).
